# Evaluating an epidemiologically motivated surrogate model of a multi-model ensemble

**DOI:** 10.1101/2022.10.12.22280917

**Authors:** Sam Abbott, Katharine Sherratt, Nikos Bosse, Hugo Gruson, Johannes Bracher, Sebastian Funk

**Affiliations:** The Centre for Mathematical Modelling of Infectious Diseases, London School of Hygiene & Tropical Medicine, London, UK; Department of Infectious Disease Epidemiology, London School of Hygiene & Tropical Medicine, London, UK; Chair of Statistical Methods and Econometrics, Karlsruhe Institute of Technology (KIT), Karlsruhe, Germany

## Abstract

Multi-model and multi-team ensemble forecasts have become widely used to generate reliable short-term predictions of infectious disease spread. Notably, various public health agencies have used them to leverage academic disease modelling during the COVID-19 pandemic. However, ensemble forecasts are difficult to interpret and require extensive effort from numerous participating groups as well as a coordination team. In other fields, resource usage has been reduced by training simplified models that reproduce some of the observed behaviour of more complex models. Here we used observations of the behaviour of the European COVID-19 Forecast Hub ensemble combined with our own forecasting experience to identify a set of properties present in current ensemble forecasts. We then developed a parsimonious forecast model intending to mirror these properties. We assess forecasts generated from this model in real time over six months (the 15th of January 2022 to the 19th of July 2022) and for multiple European countries. We focused on forecasts of cases one to four weeks ahead and compared them to those by the European forecast hub ensemble. We find that the surrogate model behaves qualitatively similarly to the ensemble in many instances, though with increased uncertainty and poorer performance around periods of peak incidence (as measured by the Weighted Interval Score). The performance differences, however, seem to be partially due to a subset of time points, and the proposed model appears better probabilistically calibrated than the ensemble. We conclude that our simplified forecast model may have captured some of the dynamics of the hub ensemble, but more work is needed to understand the implicit epidemiological model that it represents.

## INTRODUCTION

Multi-model and multi-team ensembles have become increasingly popular as an approach to increase the robustness and performance of infectious disease forecasts over the last decade (Reich et al. 2022). The experience of other domains has inspired these approaches, for example, climate modelling (IPCC, n.d.), where ensembles of both multiple models and from multiple teams have a long history of providing forecasts that stakeholders trust. The trend towards large-scale multi-team ensemble forecasting in infectious diseases has accelerated during the COVID-19 pandemic due to a pressing need for reliable forecasts and a perception that many publicly available forecasts were low quality. Over 2020 and 2021, teams established COVID-19 Forecasting Hubs covering the US (Cramer et al. 2022), Germany and Poland (J. Bracher et al. 2021), and Europe (Sherratt et al. 2022) (all three including authors of this study). All of these collaborations ensembled contributions from multiple independent teams using a similar approach and have shown that their ensemble forecasts outperform most individually contributed forecasts whilst remaining generally robust to outliers in reporting. Both the US and European Forecast Hubs were supported and received funding from public health agencies (the Center for Disease Control, CDC, and European Center for Disease Prevention and Control, ECDC, respectively) with their forecasts used in official communications by these agencies.

Whilst there is robust and consistent evidence that multi-team ensemble forecasts provide reliable and performant forecasts across domains (Reich et al. 2022) they also have a range of downsides. The most significant is the difficulty in interpreting them. This relates both to the underlying mechanisms for the forecasts they produce and to understanding if and when their behaviour is desirable. This impacts users’ trust, how easily ensemble performance can be improved, and how easily contributor forecasts can be improved. Forecasts from these ensembles also require considerable resource cost to produce as they typically require contributions from multiple independent teams, the development of several models, and a centralised group to run the ensembling project. Additional challenges with maintaining multi-team collaborations can include providing detailed feedback to those contributing forecasts that would allow them to improve their forecast approaches, providing incentives for forecasters to continue to contribute and adjust their models to changing conditions, and difficulty improving the quality of the ensemble by learning from past predictive performance (Sherratt et al. 2022). Each of these issues may impact the long-term quality of the resulting forecasts and have implications for end-users. Little progress has so far been made in mitigating these downsides or in improving access to the high-quality and robust forecasts they seek to generate for geographies without coverage or for other infectious diseases. There has also been limited critical feedback on the structure of forecasting ensembling projects for infectious disease epidemiology and little evaluation of the effort required to produce them relative to their benefits for improving forecast performance.

In climate forecasting (Castelletti et al. 2012; Edwards et al. 2021; Williamson et al. 2013), as well as in other fields such as astrophysics (Vernon, Goldstein, and Bower 2014), emulation approaches have been used to circumvent resource requirement issues for complex models by training a simplified model, usually, a non-parametric statistical model, to replicate the behaviour of either the entire model or sub-components. These approaches generally take the same inputs as the models they seek to emulate and then are trained based on the output from those models. In the context of epidemiological models, non-parametric emulation has been used to allow the rapid exploration of the parameter space of complex models that would otherwise be resource-prohibitive (Iskauskas et al. 2022; Charles et al. 2022). These methods may be less useful for resolving some of the issues of multi-team and multi-model forecasts as they do not provide interpretability, key for stakeholder take-up. Additionally, it is not clear how these methods perform out of sample, or how they would be applied to a quantile-based forecast.

In this work, we draw insights from ensemble forecasts produced and endorsed by the COVID-19 Forecast Hubs, as well as our forecasting work, to propose and evaluate a “surrogate” forecast model. This seeks to reproduce ensemble performance by mimicking its behaviour based on a minimal set of easily communicated and epidemiologically justifiable assumptions, and limited computational resources with an easily generalised implementation. The primary aim of this approach is to help highlight the behaviour, and potential mechanisms behind this behaviour, of ensemble forecasts widely considered the gold standard for COVID-19 forecasting. Our secondary aim is to provide the basis for a robust forecasting system that others can easily reuse both in operational contexts and as a platform for future research.

To achieve these aims, we evaluate an initial attempt at developing a surrogate model to replicate the observed behaviour of current multi-team forecast ensembles based on a set of clear assumptions. We submitted this model to the European Forecast Hub and here we evaluate its performance relative to the Hub ensemble. In this work, we first define the model and summarise its implementation, with a focus on minimal resource use and reproducibility as a GitHub Actions workflow (“About GitHub-hosted Runners” 2022).

We then evaluate its real-time performance in comparison to the European Forecast Hub ensemble by visualising forecasts, using the weighted interval score (Bracher et al. 2021), a commonly used proper scoring rule, and quantifying the empirical coverage of the forecasts produced. We attempt to highlight settings where this model performs well as a surrogate to the ensemble forecast and areas where it performs less well. Finally, we summarise our findings, discuss their implications, and highlight areas where more work is needed. We aim for this work to highlight some of the potential implicit assumptions of current COVID-19 Forecast Hub ensembles, provide a sensible, low-resource, surrogate model where large-scale collaborative forecasting efforts are not possible, and provide inspiration for forecasters looking to make principled improvements to their models.

## MATERIALS AND METHODS

### Setting of the European COVID-19 Forecast Hub

To understand the behaviour of the Forecast Hub ensembles we need to first explore the structure of the COVID-19 Forecast Hubs (Cramer et al. 2022; J. Bracher et al. 2021; Sherratt et al. 2022). These collaborations share a similar design with a central team running the hub, vetting forecasts, and producing the ensemble forecast as well as teams of independent forecast contributors who design their forecast models and then use them to produce a weekly forecast that they then submit to the central hub team. Each hub targets a range of metrics, including test-positive reported cases, reported deaths, and hospitalisations; has a specific geographic focus, and asks for weekly forecasts (using MMWR epidemiological weeks i.e. Sunday to Saturday (Department of Health, n.d.)) over a time horizon of a few weeks. Observed data are available and updated daily, and most submitted forecasts use this dataset, along with potentially other sources of real-time information, to produce forecasts. Here, we focus on reported cases and primarily on the European Forecast Hub but our observations hold, in our view, across COVID-19 Forecast Hubs and to a lesser degree targets. We focus on reported cases as these represent the most common forecast target for COVID-19 forecast models (Nixon et al. 2022), they are often of the most direct interest due to being a leading indicator for other metrics such as hospitalisations (Meakin et al. 2022), and they are generally the most challenging to predict (Sherratt et al. 2022). In general, 5 main classes of forecast models are submitted (Bracher et al. 2022; Cramer et al. 2022), statistical forecasting models such as ARIMA models, mechanistic forecasting models based on the compartmental modelling framework and its generalisations (Srivastava, Xu, and Prasanna 2020; Li et al. 2021), semi-mechanistic approaches that blend both of these approaches (Castro et al. 2021; Bosse et al. 2022), agent-based simulation models (Rakowski et al. 2010; Adamik et al. 2020), and human insight based forecast models that may also include elements of other methods (Karlen 2020; Bosse et al. 2022). Real-time evaluation has shown that each of these classes of models may perform well depending on the context and specific implementation of the forecast model (Bosse et al. 2022).

We extracted forecasts and data on notified weekly COVID-19 cases from the European forecasting hub (Sherratt et al. 2022; E. C.-1. F. H. Team 2021) from the 15th of January 2022 to the 19th of July 2022 for the ensemble model (referred to as the `EuroCOVIDhub-ensemble` by the hub team) and the surrogate model (submitted as `epiforecasts-weeklygrowth` and defined in the following section). We included all locations covered by the European forecasting hub which were 32 European countries, including all countries of the European Union and European Free Trade Area, and the United Kingdom. Data on notified weekly cases was originally sourced from the Johns Hopkins University (JHU) curated data repository (Dong, Du, and Gardner 2020). We used the latest available observed data as of the 1st of September 2022 (commit `f6922c3e4bdcb055abcbba8e73472afacac4cf40` from (Team 2022)). Incidence was aggregated by epidemiological week (i.e. Sunday through Saturday). As observations are subject to revisions this means that the data used to produce forecasts for a given date may not reflect the data used for evaluation. To account for this we followed the practice of the European forecasting hub project in excluding forecasts made using anomalous truth data in the week of the forecasts production and excluding forecasts for target weeks with anomalous data (Sherratt et al. 2022). We defined anomalous data based on the implementation used by (E. C.-1. F. H. Team 2021) where a data point is considered anomalous if a future revision alters it by more than 5%.

The European Forecast Hub requests forecasts for one to four-week forecast horizon and requires forecasts to use a pre-specified format with 23 quantiles of the predictive probability distribution. No restrictions were placed on who could submit forecasts and the hub team actively invited participation from research groups known to be involved with COVID-19 forecasting projects. Teams submitted forecasts at the latest two days after the complete dataset for the forecast week became available and were allowed to use all data available at the time of submission (i.e including up to two days of data for the current week). The ensemble forecast was constructed by taking the median of all forecasts for each predictive quantile without the exclusion of any validly submitted forecast (where validity was defined as passing minimal formatting checks by the hub team and timely submission) (Sherratt et al. 2022). An ensemble was only produced for locations with at least 3 independent forecast models including the hub baseline model. Submitted forecasts and target observations are available from the European Forecast Hub GitHub repository (Team 2022). We provide code in the repository of this study to streamline access.

### Observations based on previous forecasts

We have contributed a range of forecasts to COVID-19 forecast collaborations, generally focused on semi-mechanistic statistical methods and human insight-based forecasts. Our forecast submissions have not systematically over- or under-performed other forecasts submitted to the forecasting hubs (see `epiforecast` tagged models at (E. C.-1. F. H. Team 2021) and (Bosse et al. 2022; Cramer et al. 2022; J. Bracher et al. 2021; Sherratt et al. 2022)). The model-based forecasts we have contributed have focussed on trying to carefully model the underlying infectious disease dynamics from infection through to symptom onset, and test positivity using non-exponential delay distributions whilst also attempting to model the complexity of daily, within the week, reporting periodicity (Bosse et al. 2022; Abbott, Hellewell, Thompson, et al. 2020; Abbott, Hellewell, Sherratt, et al. 2020). Based on our observations our forecasts have generally captured the current trend relatively well but have not been robust to reporting issues such as large outliers in reporting and changes to reporting patterns. Our previous methodology also requires significant computational resources, running for an hour on a Azure D v5-series 16-core machine, when producing forecasts for the European forecasting hub (“Pricing - Linux Virtual Machines” 2022). This resource usage is likely beyond the capacity of many interested in having access to state-of-the-art short-term forecasts of infectious diseases. In our model-based forecasts, we did not attempt to capture potential future interventions or known interventions not currently observed in the epidemiological data whereas in our human insight models these were implicitly included. We found that our human insight-based forecasts outperformed our model-based forecasts on average. This was particularly the case when forecasting cases and at longer forecast horizons. We hypothesised that this may have been driven by including additional information not observed in the epidemiological data (Bosse et al. 2022).

Unlike our epidemiologically motivated forecast submissions, the hub ensemble forecasts were typically robust to daily reporting artefacts. They also demonstrated some ability to forecast future changes in trends that were not present in the observed data similarly to our human insight forecasts indicating the likely inclusion of either human insight, or assumptions about future interventions. In comparison to our submitted forecasts, the ensemble forecasts were less reactive to changes in trend such as from stable or reducing case incidence to increasing incidence. On the other hand, this also meant that the ensemble was less likely to adopt short-term changes in incidence and hence produced better long-term forecasts. Finally, the ensemble forecast tended to produce sharper forecasts and have a tendency toward under-vs overpredicting. Our observations are summarised in Table 1.

**Table 1.**
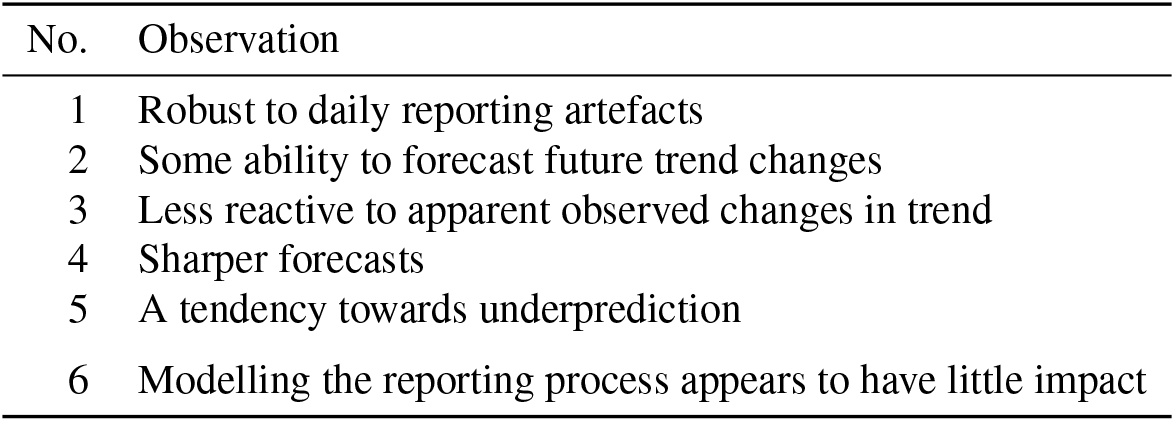
Observations on the relative performance of the Forecast Hub ensemble compared to our forecast submissions.

| No. | Observation |
| --- | --- |
| 1 | Robust to daily reporting artefacts |
| 2 | Some ability to forecast future trend changes |
| 3 | Less reactive to apparent observed changes in trend |
| 4 | Sharper forecasts |
| 5 | A tendency towards underprediction |
| 6 | Modelling the reporting process appears to have little impact |

### Model

#### Assumptions and simplifications

Based on our observations of forecast performance (summarised in Table 1), here we define a model with similar, but simplified, epidemiological characteristics to our previous approaches to model-based forecasting (Bosse et al. 2022) to produce an ensemble-like performance without sacrificing interpretability and with a lower cost to produce. The first simplification we make is to model only weekly data, rather than using daily data and then aggregating. This mitigates the impact of daily reporting artefacts. It also serves to increase the auto-correlation of the forecasting model as there is an increased lag before changes in daily observations gain significant weight in the model. This leads to the observed ensemble behaviour of being relatively auto-correlated and resistant to short-term changes in trend.

The second simplification we make is to ignore the underlying latent infection process and focus only on the observed reported cases. This removes the need for, potentially misspecified, external information on the delay from infection to report, and reduces computational requirements due to a reduction in model complexity. However, this sacrifices some of the interpretability of the forecast model as any transmission statistics we now calculate will be based on reported cases and not latent infections. As discussed in (Gostic et al. 2020) this leads to varying amounts of bias depending on the epidemic phase.

The final simplification is to model the growth rate as a differenced auto-regressive process with an order 1 rather than using a gaussian process-based method as we have done in other work (Bosse et al. 2022; Abbott, Hellewell, Thompson, et al. 2020; Abbott, Hellewell, Sherratt, et al. 2020). This represents a parsimonious approach in that we encode our expectation that the growth rate should vary over time and allow this to influence the forecast but we include only a single lag term, reducing the computational overhead of the model. To model potential unobserved interventions and more general changes in transmission, we include an additional growth rate modifier restricted to be between 0 and 1 that differs depending on if the growth rate is positive or negative (due to potential differing responses when cases are growing or increasing) and that acts in a multiplicative fashion (meaning that larger absolute growth rates are reduced to zero growth more rapidly). This reflects a simplified interpretation of how the ensemble appears to react to potential future changes by assuming a gradual return to stable incidence.

The only observation for which we do not make an adaptation is the apparent sharpness of the ensemble compared to our prior forecasting models. Instead, we make use of a negative binomial observation model allowing the inclusion of overdispersion. This choice is motivated by our belief that the underlying transmission process is an exponential discrete one and therefore a count error model with a log link function, where variance is linked to the mean, is a sensible choice. We suggest that part of the reason the hub ensembles exhibit such sharpness is due to the penalisation of overprediction compared to underprediction caused by the use of a generalised form of absolute error for the majority of forecast evaluations (Bracher et al. 2021). Our set of assumptions and simplifications are summarised in Table 2.

**Table 2.** Assumptions/simplifications based on observations of the relative performance of Forecast Hub ensembles compared to our forecast submissions.

| Assumption | Observation |
| --- | --- |
| Reported cases can be modelled using weekly data and a generative process discretised by week | 1, and 2 |
| Reported cases can be modelled as if they represented infections | 6 |
| The growth rate of infections can be represented as an auto-regressive process with an order of 1 week | 3 and 4 |
| Unobserved interventions and more general changes in transmission towards a stable state can be represented using a multiplicative decay parameter | 2, and 5 |

##### Definition

We model the expectation (*λ*_*t*_) of reported cases (*C*_*t*_) given past reported cases as an order 1 autoregressive (AR(1)) process by epidemiological week (*t*) on the log scale. The model is initialised by assuming that the initially reported cases are representative with a small amount of error (2.5%). We assume a negative binomial observation model with overdispersion *ϕ* for reported cases (*C*_*t*_).

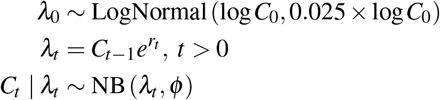

where the mean and variance of the negative binomial are given by

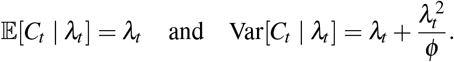

Here *r*_*t*_ can be interpreted as the weekly growth rate. *r*_*t*_ is then modelled as a piecewise constant differenced AR(1) process modified such that the dependence of *r*_*t*-1_ is multiplied by a decay factor (*ξ*_+,−_) that varies dynamically according to the sign of *r*_*t*-1_. This assumes that the growth rate is non-stationary with a trend that is independent of the current growth rate (the differenced AR(1) process), the additional decay factor encodes the belief that larger absolute growth rates will tend more quickly towards no growth and that this process may work differently for positive or negative growth rates. This process can be defined as follows,

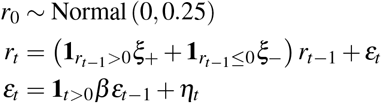

where *ε*_*t*_ and *η*_*t*_ are error terms. The following priors are used,

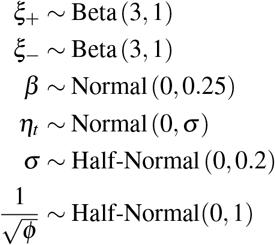

Where *σ*, and 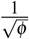 are truncated to be greater than 0 and *β* is truncated to be between -1 and 1. The Beta priors for *ξ*_+,−_ have been chosen to be weakly informative that the reduction towards 0 growth is relatively slow. Similarly the prior for *β* has been chosen to be weakly informative that there is weak auto-correlation in differenced growth rates. *σ* has also been made weakly informative under the assumption that the potential change in growth rates in a single time-step should be relatively small.

### Forecast evaluation

We standardised the magnitude of observations and forecasts across forecast locations, in order to facilitate comparison, by scaling both weekly notified test positive cases and forecast test positive cases by the population in the forecast region to an incidence rate per 10,000 people. This differs from the approach typically taken by the Forecast Hubs where no population standardisation is used (Cramer et al. 2022; J. Bracher et al. 2021; Sherratt et al. 2022). We then visually evaluated forecasts from a subset of locations by forecast horizon (1 and 4 weeks) on both the natural and log scales. The countries in this subset were Germany, Greece, Italy, Poland, Slovakia, and the United Kingdom. These countries were selected to include forecasts based on different numbers and types of submitted forecast models, to be at least partially representative of the full sample of forecast locations, and to include nations for which the authors had a good understanding of local data and transmission dynamics in the study period.

We evaluate forecasts for all locations and horizons quantitatively using the absolute error (AE) of the median forecast and the weighted interval score (WIS) (Bracher et al. 2021). The WIS is a quantile-based proper scoring rule that approximates the continuous ranked probability score (CRPS). Both the WIS and CRPS are generalisations of the absolute error to evaluate probabilistic forecasts and are widely used to evaluate COVID-19 forecasts, including by the European Forecast Hub (Sherratt et al. 2022). We present WIS for the subset of forecasts we explore visually for both the ensemble and surrogate model by date and forecast horizon (1 and 4 weeks).

To understand the relative performance of the surrogate model compared to the ensemble model, we calculate the relative performance (rWIS and rAE) by dividing the WIS/AE for the surrogate model by the WIS/AE of the ensemble model for all locations and forecast horizons. To maintain the propriety of this score, we do this after first taking the means of scores for the relevant stratification. We explore relative performance by forecast horizon, by month and horizon, and by location and horizon.

In addition to presenting the WIS for a subset of locations and the relative WIS for all locations, we also calculate and visualise the empirical coverage, which is the percentage of observed values within a given interval or below a given quantile, of both the surrogate and ensemble model for the 30%, 60%, and 90% prediction intervals and by quantile (N. I. Bosse et al. 2022). We also calculate the bias (see (N. I. Bosse et al. 2022) and (Funk et al. 2019) for a more detailed definition) of both forecasting approaches, stratified by forecast horizon. This metric aims to capture the tendency for a forecast to under or over-predict. It captures the average proportion of the mass of the forecast distribution that is above or below the true value (and so can range from -1 to 1) with an unbiased forecast having an average bias value of 0. Lastly, we calculate and visualise the relative weighted interval score by quantile, stratified by forecast horizon, to assess the relative difference in performance across the predictive distribution.

### Implementation

The model is implemented in `stan` (S. D. Team 2021) and `R` (`4.2.0`) (R Core Team 2019) as an extension of the baseline model from the `forecast.vocs` R package (`0.0.9.7000`) (Abbott 2021). We note that our use of an indicator function introduces a discontinuity to the posterior making it less suited for use with `stan`. Other model formulations without this feature would be more efficient and robust. The `cmdstanr` R package (`0.5.2`) (Gabry and Češnovar 2021) is used for model fitting with 2 MCMC chains each having 1000 warm-up and 1000 sampling steps each (Gabry and Češnovar 2021). `cmdstanr` surfaces several settings that trade-off between sampling speed and the robustness of the approach. Here we take a conservative approach, as the model fit is not manually inspected during real-time usage and due to the expected complexity of the posterior (Betancourt 2017), and set the adapt delta setting to 0.99, and the maximum tree depth setting to 15. For real-time usage, convergence was not assessed, but during model development, the Rhat diagnostic was used alongside feedback from `cmdstanr` about the number of divergent transitions and exceedance of the maximum tree depth (Gabry and Češnovar 2021). During development, posterior predictions were also visually compared to observed data.

To download and manipulate forecasts from the European forecasting hub (E. C.-1. F. H. Team 2021) we use the `data.table` (`1.14.2`) (Dowle and Srinivasan 2021) and `gh` (`1.3.0`) (Bryan and Wickham 2021) R packages. We make use of further functionality from the `forecast.vocs` R package (Abbott 2021) to prepare data for forecasting, visualise forecasts and summary measures, and summarise forecasts. Forecast evaluation is implemented using the `scoringutils` R package (1.0.0) (N. I. Bosse et al. 2022), and the `scoringRules` R package (1.0.1) (Jordan, Krüger, and Lerch 2019).

To ensure the reproducibility of this analysis dependencies are managed using the `renv` R package (`0.14.0`) (Ushey 2021) and a Dockerfile file along with a built Docker image (Boettiger 2015) (via GitHub Actions (“About GitHub-hosted Runners” 2022)) is provided in the code repository. Weekly forecasts were made using `renv` and based on GitHub Actions free tier as available in 2022 to ensure they require limited compute and that our implementation is independent of local resources facilitating democratised access. The free GitHub Actions runner we used for all forecasts was Ubuntu `20.04` based with 2 cores (x86 64), 7 GB of RAM, and 14 GB of SSD space. The code for this analysis can be found here: https://github.com/epiforecasts/simplified-forecaster-evaluation The code for the forecasting model defined above along with the infrastructure required to forecast using GitHub Actions can be found here: https://github.com/seabbs/ecdc-weekly-growth-forecasts Versions archived on Zenodo are available (Abbott and Bosse 2022) and (Abbott and Sherratt 2022).

## RESULTS

### Summary of the European COVID-19 Forecast Hub Setting

In our study period, incidence rates across European nations and in the UK were primarily driven by the spread of novel subvariants of concern related to the Omicron variant and changes in population susceptibility. Many countries, such as the UK, saw large BA.1 waves in January, resulting in declining incidence rates through February (Figure 1). From late February through to the end of May, most nations saw another wave driven by BA.2. This wave typically saw lower reported incidence rates, and was characterised by a lower peak than the BA.1 wave with a more gradual decrease in incidence. The end of our study period was dominated by the gradual take-over of the BA.4/BA.5 subvariants that again had a lower peak and lower absolute growth rates. Unlike earlier periods in the pandemic, our study period did not see the use of new non-pharmaceutical interventions (NPIs) in response to increasing COVID-19 incidence in most locations. In addition, ascertainment rates likely reduced over time in most locations due to reductions in routine testing and test availability. Whilst both the reduced use of NPIs and testing generally occurred across nations our study period also marked an increase in the heterogeneity of the response to the COVID-19 pandemic with nations changing policy at different times and to different degrees. This is in contrast to the early COVID-19 pandemic response for which most nations took similar actions at similar times.

**Figure 1.**
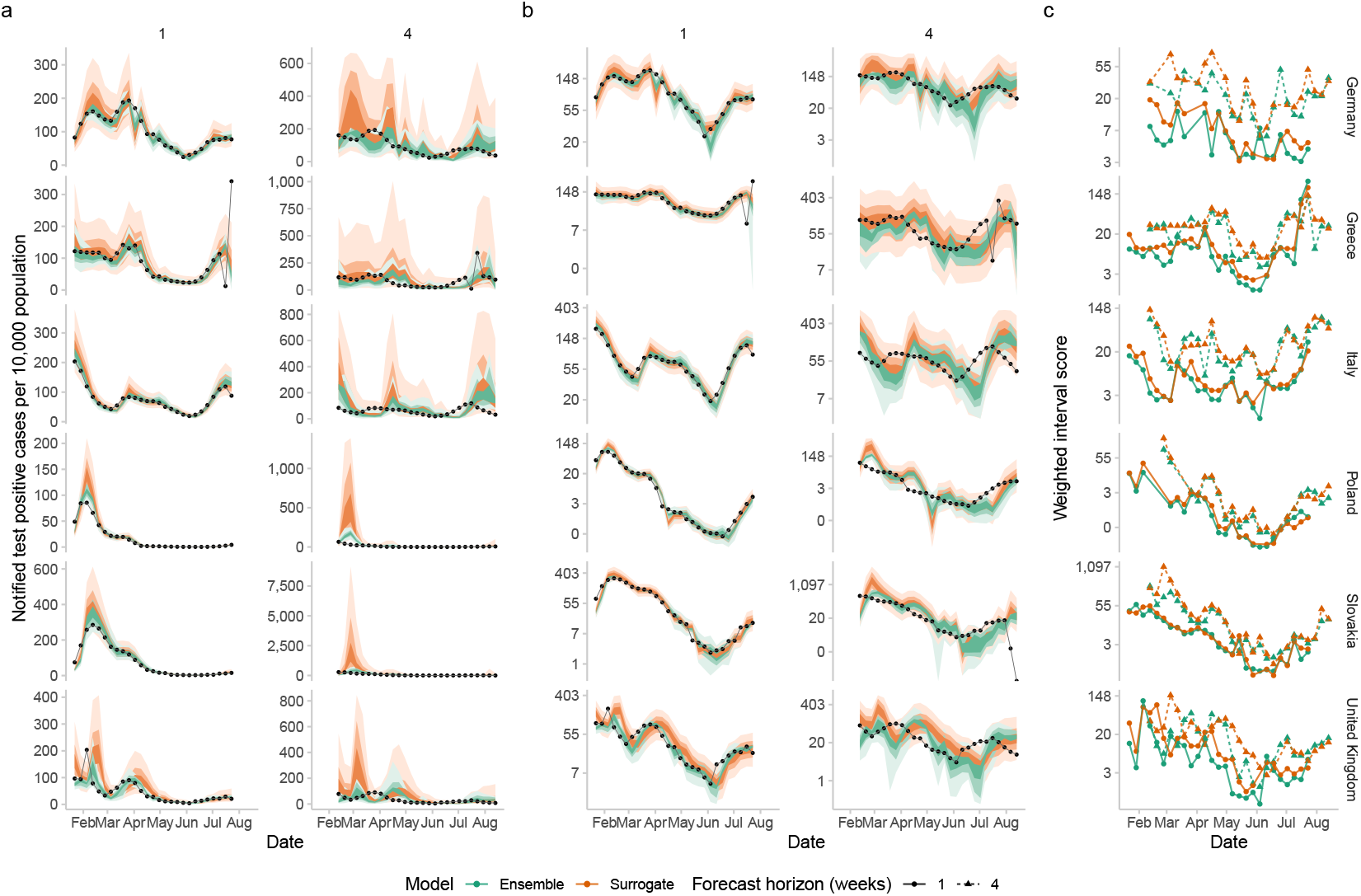
a.) Forecasts of notified test-positive cases (per 10,000 population) by epidemiological week in Germany, Greece, Italy, Poland, Slovakia, and the United Kingdom, by forecast horizon (one and four weeks) for the surrogate model (orange) and forecast ensemble (green). 30%, 60%, and 90% prediction intervals are shown. The black line and points are the notified cases as of the date of data extraction rather than those available at the time. b.) A replicate of a.) but with incidence rates on the log scale. c.) Weighted interval scores at the one-week and four-week forecast horizon by epidemiological week in Germany, Greece, Italy, Poland, Slovakia, and the United Kingdom on the log scale.

We extracted forecasts starting from the 15th of January until the 19th of July 2022 for all countries covered by the European forecasting hub (nations of the European Union, the European Free Trade Agreement, and the United Kingdom, making 32 unique locations). In total 8846 forecasts were made across all locations, with 27 unique forecast dates and 32 independent forecast models (including the European hub baseline model). Of these models, 10 forecasted in at least 30 locations including our original submission (referred to as `epiforecasts-EpiNow2` by the hub), and our surrogate model. Of the remaining models submitted 16 were submitted in only one location. Single-location models were clustered in a few locations, particularly in Germany and Poland (likely due to the folding of the German/Poland forecasting hub into the European forecasting hub project (Sherratt et al. 2022)). Italy was also an outlier with 4 models that submitted nowhere else. 4 models were submitted for between 3 and 30 locations and all these models varied the number of locations they submitted forecasts for over time, potentially indicating manual curation or models targeted at specific conditions.

Across all forecast dates and locations the minimum number of independent forecasts was 4 with the maximum being 20. The median number of independent forecasts per location and forecast date was 10. All locations received forecasts from at least 10 models with the median number of forecast models per location being 12. Coverage of forecast dates varied across submitted models with 8 models submitting for all dates, 16 models submitting for at least 90% of dates, and 6 models submitting for fewer than 50% of forecast dates. In general, there was no clear difference in forecast date coverage between models that submitted for all locations vs a small subset but models with partial coverage of locations all also had partial coverage of forecast dates.

63 observations, stratified by week and location, were defined to be anomalous within the study period by the European Forecast Hub (E. C.-1. F. H. Team 2021). Forecasts for these observations were excluded as were forecasts for forecast weeks where they were the latest available data. Data anomalies were not randomly distributed with some locations being particularly prone to data revisions including Lithuania (with 23 weeks with data anomalies), and Portugal (with 13 weeks with data anomalies). Anomalies were also not evenly distributed over time with a higher proportion occurring earlier in the study period (potentially due to our choice to extract data from the 1st of September which effectively truncated anomalies). 7.3% of forecasts were excluded across all horizons due to anomalies in the observed data. Aggregated across horizons 10.3% of forecasts included at least one week with anomalous data.

### Forecast evaluation

#### Visualisation of forecasts by horizon

In our example set of locations, the absolute performance of the ensemble and the surrogate model was visually similar on the log scale in all locations at short forecast horizons though this varied by location (Figure 1 b). On the natural scale the difference in performance was more marked, especially for periods of peak incidence and at longer horizons (Figure 1 a). Performance was not homogeneous across our set of example locations with the surrogate model performing similarly to the ensemble in Slovakia whilst in the United Kingdom and Germany the surrogate model performed substantially worse for some forecast dates (Figure 1). For both the ensemble and the surrogate, performance decreased as the forecast horizon increased with this being particularly noticeable for the surrogate model during periods of peak incidence. In general, in the study period, the ensemble appeared to be better able to forecast peak incidence. Both models forecast large reductions in incidence in Poland during May that did not occur whilst only the ensemble forecast spuriously forecast similar large reductions in Germany during June. In comparison to the ensemble model the surrogate model appeared less likely to place weight on unfeasibly large reductions in incidence during periods of declining incidence but on other hand was more likely to forecast continuing increases in incidence (for example in February in Slovakia and Poland).

#### Relative forecast evaluation

Evaluating the ensemble and surrogate models using the WIS across all locations and forecast dates we found that the mean relative performance of the surrogate model was 1.27 at the one-week horizon, 1.28 at the two-week horizon, 1.4 at the three-week horizon, and 1.69 at the four-week horizon, indicating that the ensemble forecast outperformed the surrogate forecast for all horizons by at least 25% and that the relative performance of the surrogate model degraded as the forecast horizon increased (Figure 2 c). Much of this outperformance, especially at longer forecast horizons, was driven by a small subset of forecasts with relative performance having a heavy tail (Figure 2 a). If we instead consider median relative performance (note this is not a proper scoring rule and should not be used to choose between models) we find that, relative to the ensemble, the surrogate scored 1.21 at the one week horizon, 1.14 at the two week horizon, 1.2 at the three week horizon, and 1.28 at the four week horizon. This would suggest that an increasingly skewed score distribution as the forecast horizon increased is responsible for the increase in the mean relative score (Figure 2 a). 31% of individual surrogate forecasts scored better than the comparable ensemble forecast, 68% performed within 50% of the comparable ensemble forecast, and 17% had a more than 100% worse WIS than the comparable ensemble forecast.

**Figure 2.**
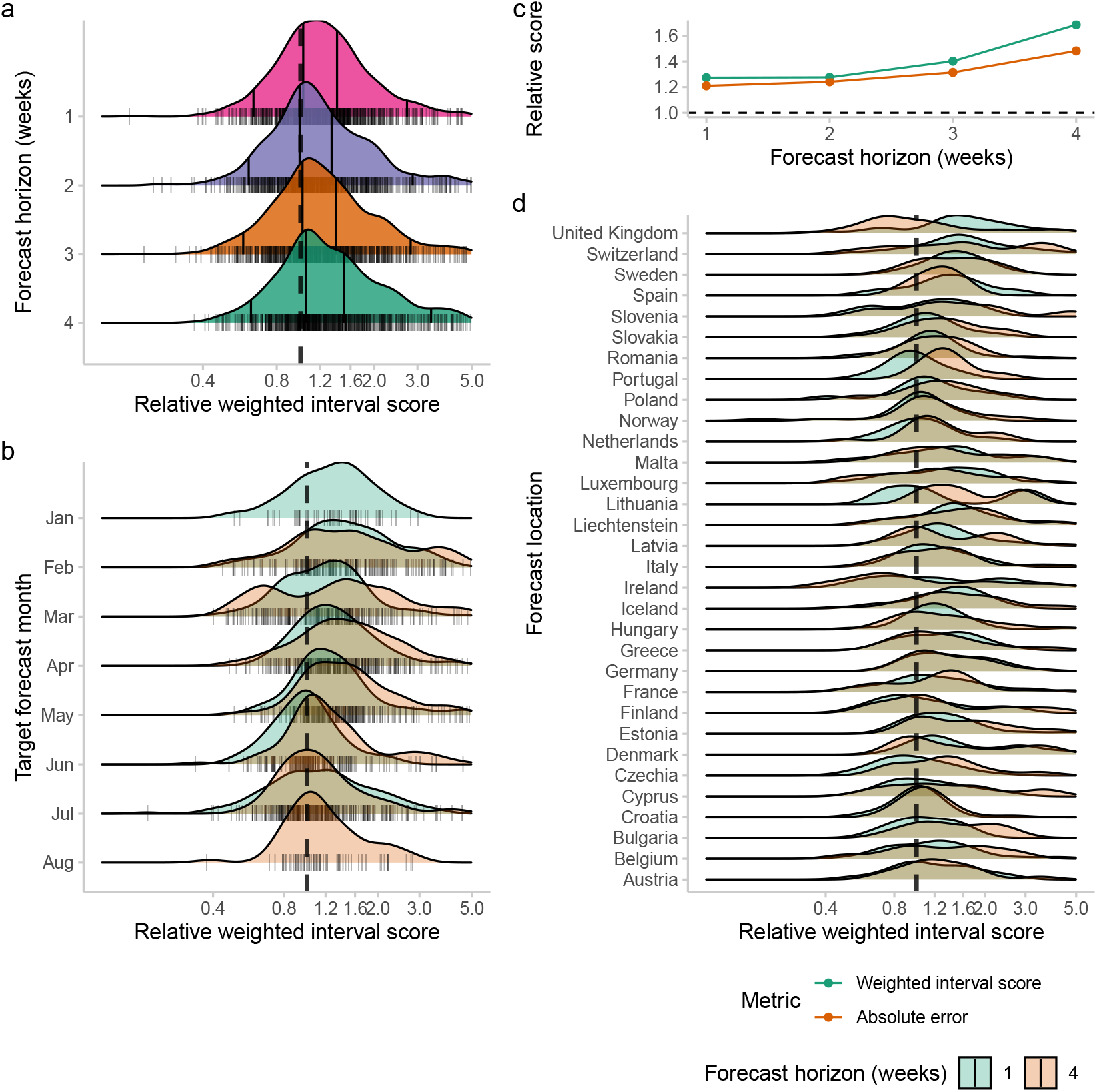
Relative weighted interval score by location, horizon, and forecast date for the surrogate forecast model compared to the ensemble forecast model on the log scale. a.) The density of the relative score by horizon. Horizontal black lines give the 5%, 35%, 65%, and 95% quantiles. b.) The density of the relative score by month for a given forecast horizon stratified by the one and four-week forecast horizon. c.) The average relative weighted interval score and absolute error for the surrogate model compared to the ensemble forecast by forecast horizon. d.) The density of the relative score by forecast location stratified by the one and four-week forecast horizon. The dashed line on all plots indicates when the ensemble forecast is equivalent to the surrogate forecast. The vertical black lines on the y-axis give individual relative scores.

If we consider only the median point forecast, using the absolute error, we see that the ensemble forecast again outperformed the surrogate forecast (rAE for the surrogate compared to the ensemble 1.34). If we instead consider the median of the absolute error we see that the difference in performance has reduced indicating a similar skewed score distribution for point forecasts as for the whole predictive distribution (rAE 1.11). Across forecast horizons the same pattern of outperformance holds. However, the difference in relative performance was less than when the full probability distribution was accounted for, with this becoming more marked as the forecast horizon increased (Figure 2 c).

The surrogate model’s relative performance varied over time with substantially worse performance from January to March compared to later in the year across all forecast horizons based on changes in the relative score distribution and its summary statistics (Figure 2 b). The majority of the difference in performance appeared to be driven by a thicker right tail with this being a particular feature of forecasts at longer horizons. Forecast performance in March had a bimodal distribution at the four-week horizon with a substantial fraction of surrogate forecasts outperforming the ensemble and a substantial fraction substantially underperforming. This variation in performance may have been linked to the BA.2 wave which peaked in most locations during this period if the surrogate model was more likely to overpredict peak incidence than the ensemble forecast.

There was also substantial variation across forecast locations with the surrogate performing relatively well in some locations at some forecast horizons, for example, the four-week horizon in the United Kingdom, and badly in others, for example, the four-week forecast in Switzerland (Figure 2 c). In general, across locations, as observed overall, relative forecast performance degraded across horizons with a heavier right tail at longer horizons. Some locations showed less of this behaviour, for example, Spain, and in some, it was very dominant, for example, Switzerland.

#### Forecast calibration

Overall the surrogate model was relatively well calibrated at the 30%, 60% and 90% prediction interval, though with a tendency to be slightly underconfident, with empirical coverage of 30.5%, 62.5%, 92.3% respectively. The ensemble model was less well calibrated, with a tendency to be overconfident with empirical coverage of 24.8%, 51%, 79% respectively (Figure 3 a). When stratified by forecast horizon the ensemble forecast was best calibrated at the one-week forecast horizon, and then became progressively less well calibrated as the forecast horizon increased (Figure 3 a). In comparison, the surrogate forecast was less well calibrated than the ensemble forecast at the one-week forecast horizon with a tendency to have a larger empirical coverage than required (Figure 3 a). At longer horizons and narrower prediction intervals, the surrogate forecast became better calibrated though with a tendency to be overconfident. This was not the case for the 90% prediction interval where the surrogate model covered more than the expected interval, for all horizons, indicating forecasts were overly uncertain for this interval regardless of the horizon.

**Figure 3.**
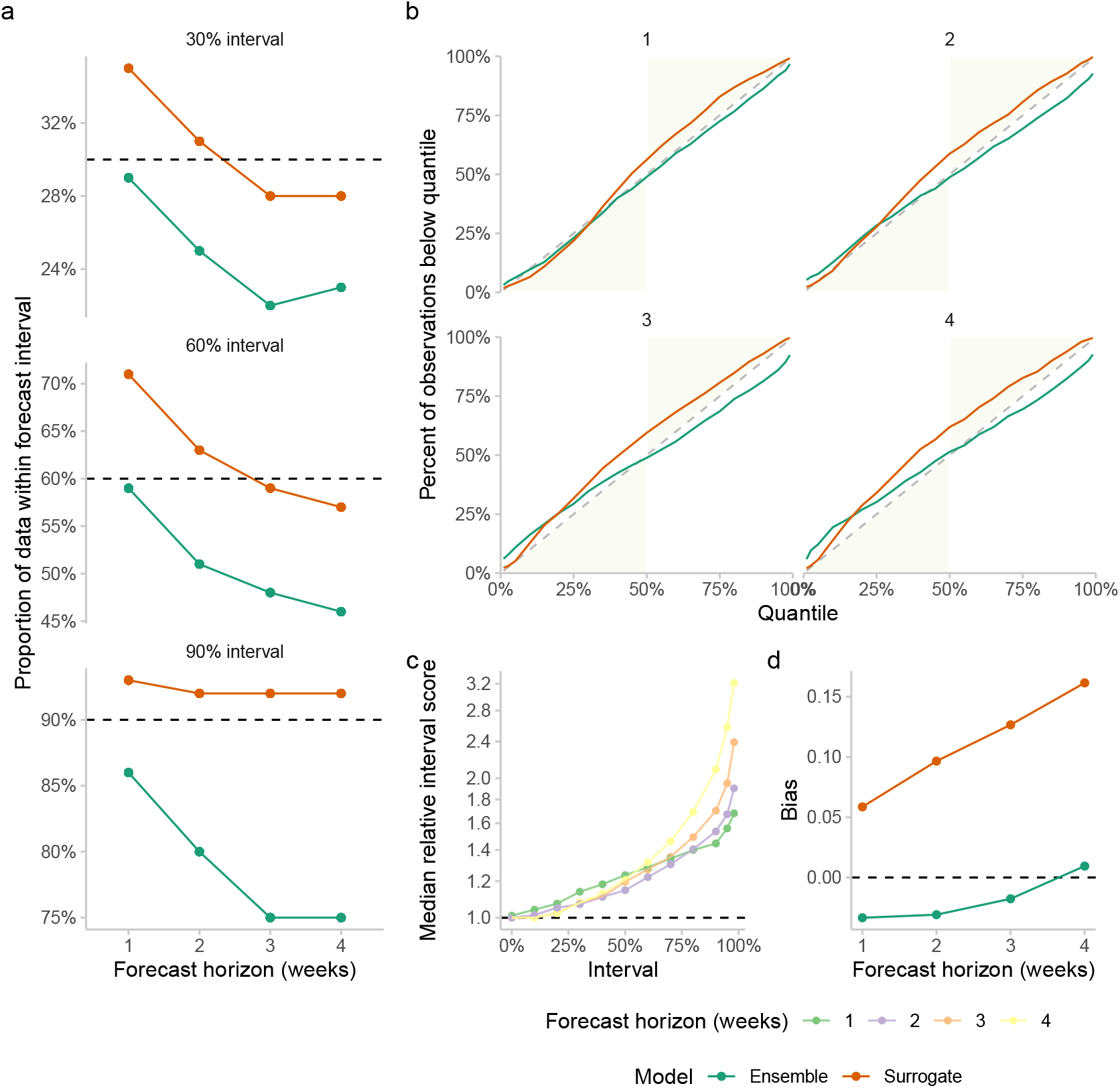
a.) Empirical coverage of the surrogate (orange) and ensemble (green) forecasts at the 90%, 60%, and 30% prediction intervals stratified by forecast horizon. Ideally, a well-calibrated forecast should have empirical coverage for a given prediction interval that equals the nominal level of the interval (i.e., 30%, 60% and 90%, respectively). b.) Empirical coverage by quantile for both the surrogate and ensemble forecasts. A well-calibrated forecast should have empirical quantiles that match the theoretical ones. The green area of this figure corresponds to conservative forecasts. c.) Median relative weighted interval score by quantile and forecast horizon for the surrogate forecast compared to the ensemble forecast. d.) The bias of the ensemble and surrogate forecasts stratified by horizon.

Stratifying calibration by quantile and forecast horizon the ensemble forecast was conservative at all horizons for quantiles larger than the median whilst being comparably well calibrated for intervals below the median (Figure 3 b). This behaviour became more prominent as the forecast horizon increased. In contrast, the surrogate forecast was generally equally well calibrated across horizons with a tendency to be under confident for intervals above the median. At longer horizons, however, quantiles below the median were over confident.

Breaking down the relative weighted interval score by forecast interval we observe that the surrogate model produces forecasts that differ most from the ensemble in the outer intervals and in particular the tails of the forecast (Figure 3 c). This is true across forecast horizons but the magnitude of the difference increases.

Calculating the bias of the forecasts from each model we see that the (Figure 3 d) ensemble forecast is initially biased towards underprediction but this bias reduces as the forecast horizon increases. In comparison, the surrogate forecast model is biased towards overprediction for all forecast horizons with the magnitude of this bias appearing to increase linearly with the forecast horizon.

## DISCUSSION

### Summary

In this study, we defined a surrogate model aiming to replicate some of the observed behaviour of the European Forecast Hub multi-team ensemble for forecasting test-positive reported COVID-19 cases in European nations. We first defined a set of assumptions for how the surrogate model should behave based on our observations of the European Forecast Hub ensemble, and our experience submitting forecasts to various Forecast Hubs. We aimed for a model that could be easily understood, that produced epidemiologically meaningful summary statistics, and that could be run with low compute resources. We further provide a fully reproducible workflow for running and evaluating this model using GitHub actions facilitating others to do the same.

Over the 6 months of the study period, we found that our surrogate model produced forecasts that were visually similar to those from the Forecast Hub ensemble on the log scale though with greater uncertainty. Visual differences were more marked on the natural scale with the surrogate model forecasting spuriously high peak incidence. In a subset of example locations, we observed some variation in performance across locations, that the ensemble better-captured peak incidence, and that the surrogate model appeared biased toward overprediction. Evaluating the relative performance of the surrogate model compared to the European Forecast Hub ensemble we found that the mean performance was substantially worse and that relative performance decreased with the forecast horizon. The median forecast performance of the surrogate model was also worse when compared to forecasts from the ensemble though the majority of surrogate forecasts were within 50% of the performance observed for the ensemble forecast. The difference in mean and median relative performance suggested a skewed distribution in scores, which we confirmed visually. This means that a relatively small fraction of forecasts were responsible for a substantial portion of the difference in performance. Evaluating point forecast performance indicated a similar pattern of performance as that observed using the full predictive distribution though the relative performance of the surrogate model generally improved. Performance varied by location and forecast date with the surrogate model performing worse in the first part of 2022 which may have been linked to incidence rates peaking across forecast locations linked to the spread of BA.2. In general, the relative performance of the surrogate model degraded as forecast horizons increased with the distribution of relative performance having an increasingly heavy right tail as the forecast horizon increased indicating a greater share of forecasts performing very poorly in comparison to the hub ensemble. The Forecast Hub ensemble was poorly calibrated, particularly at longer forecast horizons and larger prediction intervals, compared to the surrogate model though the surrogate model tended to be overly uncertain at large intervals. The ensemble forecast was biased towards under-prediction at short to medium forecast horizons but unbiased at longer horizons. In comparison, the surrogate model was biased towards overprediction and this bias increased linearly with the forecast horizon.

### Strengths and Weaknesses

Our study benefits from having been conducted using forecasts produced in real-time, rather than retrospectively, and submitted to an independent forecast research hub (though we note the overlap between authors on this study and the European Forecast Hub (Sherratt et al. 2022)). This means that our results are not subject to hindsight bias. The downside of this approach is that it was not possible to update the surrogate model over time in response to the initial evaluation or to explore other parameterisations that might be more successful of which there are likely several. However, as our study has been conducted with a focus on reproducibility and openness our findings can be replicated or extended by others regardless of compute availability (due to our use of GitHub actions as a compute platform which is freely available to researchers). An additional downside to this approach is that the hub ensemble includes forecasts from our surrogate model, increasing the similarity between the two approaches. This is difficult to avoid without retrospectively re-calculating the ensemble using the same approach as taken by the hub which would reduce the independence of the hub ensemble as a source of truth to compare our forecasts against. Given the number of forecasts submitted in most locations and the European Forecast Hubs’ practice of not calculating an ensemble when fewer than 3 independent forecasts were available, the bias in our results caused by this limitation should be relatively small. Notably in this study, we focussed on replicating the Forecast Hub ensembles’ observed behaviour rather than attempting to define an optimal forecast for forecast consumers. It is possible that if we had instead aimed to develop a forecast methodology that minimised the evaluation criteria we planned to use, especially if we relaxed our assumed compute resource constraints, we would have produced forecasts that performed better relative to the hub ensemble. However, if we start from the view that the Forecast Hub ensemble has traits that are desirable for use by policy-makers (i.e robustness and good average performance), which can be found widely in the literature (Cramer et al. 2022; J. Bracher et al. 2021; Sherratt et al. 2022), then our approach may make sense as a way of producing a “good” forecast without sacrificing interpretability.

Developing forecast methodologies with limited resources is critical as whilst improving predictive performance is a key goal of short-term forecasting it is also important that forecast models be accessible as this makes it easier to iteratively improve them, and makes them more flexible when used in real-time settings (Osthus 2022). An example of the lack of flexibility of the Forecast Hub ensemble, and its constituent models, is the ensembles response to upswings linked to variant dynamics, with the growth of one variant being temporally hidden by the decline of another. Rather than forecasting this ahead of time the Forecast ensembles generally only reacted to changes in the observed data indicating that variant information was not being used by most forecasters. Unlike the Forecast Hub ensemble the surrogate model can be, and indeed has been (Abbott, Sherratt, and Funk 2021), easily extended to capture this. Other examples where additional transient information is available to forecasters can be readily thought of implying this is a general advantage of simpler methods.

Our focus on replicating the performance of the hub ensemble is also useful as the surrogate model may highlight some of the emergent behaviour of the ensemble captured in our assumptions, such as auto-correlation across time points, and the growth rate tending towards zero as the forecast horizon increases. It also highlights some of the differences between our surrogate forecast model and the ensemble that may lead to new insights into the mechanisms leading to the ensemble’s behaviour, such as the generally poor coverage of the ensemble that could not be explained by the assumptions we used in developing our surrogate methodology. Whilst we normalised reported cases to be population-adjusted incidence rates, and so can more easily compare across locations than using the approach commonly implemented by the Forecast Hubs (Cramer et al. 2022; J. Bracher et al. 2021; Sherratt et al. 2022), our results are still conditional on the use of the weighted interval score as an evaluation metric. As this proper scoring rule scales with the order of magnitude of the predicted quantity this means that forecasts during periods of higher incidence are given more weight than forecasts from periods of low incidence. It also means that overprediction is penalised more than underprediction as incidence rates are bounded at zero but relatively weakly bounded by populations at the upper bound (as incidence rates are typically only a small fraction of the overall population). This bias could explain the relatively poor performance of the surrogate model, compared to the ensemble, despite the surrogate model being comparably well-calibrated. We considered alternative methods of forecast evaluation that would be robust to this potential source of bias but choose to stick relatively closely to the methodology used by the European Forecast Hub (Sherratt et al. 2022), aside from the use of population weighting to facilitate comparison between forecast locations, as these choices inform the development of submitted models and so are key to our findings.

### Literature context

There are no other studies in the epidemiology literature which we are aware of that attempt to develop a forecasting model based on the observed behaviour of a multi-team, multi-model ensemble. Few studies focus on delivering computationally feasible forecasting models in a reproducible framework backed by an openly accessible compute platform. However, the US (Cramer et al. 2022), European (Sherratt et al. 2022), and Germany/Poland (J. Bracher et al. 2021) forecasting hubs have published a range of evaluations of forecasts submitted to their platforms and the relative performance of their ensembles. In general, these studies have struggled to draw general conclusions about the structural assumptions of forecast models they consider “good” (generally they have defined this as minimising the weighted interval score, as in this study).

The poor calibration of the forecast ensembles produced by median Hub ensembles has been noted repeatedly (Cramer et al. 2022; J. Bracher et al. 2021; Sherratt et al. 2022) but little progress has been made in understanding the causes or suggesting alternatives. Progress in understanding which structural model features lead to better infectious disease forecasts has been limited. The US Forecast Hub identified the top 5 performing models and noted the structural assumptions they made, but couldn’t directly link assumptions with performance (Cramer et al. 2022). They also did not extensively compare and contrast these conclusions to arrive at a set of desired forecast assumptions (as done in this study to motivate the surrogate model), or explore the performance of a forecasting model designed with these assumptions in mind. Similarly, the Germany and Poland forecasting hubs were able to identify forecast models that performed comparably as well as their ensemble forecasts but did not derive structural assumptions that led to this out-performance or detail explicitly what the desirable performance characteristics would be, aside from optimising the weighted interval score. All comparable Forecast Hub projects found that their ensemble was often the best choice, had desirable characteristics such as robustness - though this was rarely fully defined - and should be the output used by forecast consumers (Cramer et al. 2022; J. Bracher et al. 2021; Sherratt et al. 2022). In general, during the study period, all projects used the same unweighted median ensemble forecast of all submissions. The US (Ray et al. 2022), and European (Sherratt et al. 2022), forecasting hub also evaluated a range of other ensemble approaches, such as inverse weighted interval score weighting, unweighted ensembles of a selection of models based on recent performance, and mean ensembling. Work on this is still ongoing but these more complex ensembling approaches were shown to outperform the median of all submitted forecasts in many cases in the case of the US forecasting hub and did not outperform in the case of the European forecasting hub. No Forecast Hub has switched to these alternative ensemble designs for their operational forecast of reported cases, though the US hub has switched to a trained ensemble for death forecasts. This suggests that the hub teams do not think the evidence base is strong enough for trained ensembles to be used by forecast consumers for reported cases and hence the median of all submitted forecasts remains the community-suggested default ensemble option and a sensible target for our study.

Other studies have been published evaluating single forecast models in comparison to ensemble performance from the Forecast Hub. In general, these have not focussed on replicating ensemble behaviour but rather optimising the target evaluation metric. Our previous work also highlighted the lack of calibration in an ensemble forecast from the Germany/Poland forecasting hub compared to forecasts from epidemiological models and noted the bias towards underprediction observed in the ensemble forecasts and not in our model-based forecasts (Bosse et al. 2022; J. Bracher et al. 2021). Finally, our results are potentially sensitive to the definition used to define anomalous observations (generally related to retrospective data revisions). Here we follow the practice of the European Forecast Hub (E. C.-1. F. H. Team 2021) of excluding forecasts for weeks with a data revision of more than 5% and forecasts made based on data that is subsequently revised by more than 5%.

### Further work

Whilst we derived our surrogate model from a range of assumptions based on observing ensemble forecasts behaviour and the behaviour and structure of submitted models avenues for future improvement remain in terms of improving the approach used to elicit these observations. In follow-up work, a more rigorous approach to this could be taken to further refine this set of assumptions, in particular using the input of a wider pool of researchers. The findings from our study may also be useful for informing this improved set of assumptions. A particular focus should be on understanding why our surrogate model was liable to overestimate peak incidence and what simple additional assumptions may be used to mitigate this. In addition, the model we derived based on our assumptions was likely not optimal both in terms of compute time and accuracy at reproducing ensemble-like behaviour. Models with a more complex auto-correlation structure and more refined approaches to localised trends should be explored to improve relative performance to ensemble forecasts. An example of a family of possible approaches are structural time series models which have many of the characteristics implied by our assumptions for how forecast ensembles typically operate. As we identified that the tails of our predictive distributions were responsible for a large proportion of the difference in performance compared to the forecast ensemble it may be the case that post-processing of forecasts from our surrogate model would enhance their similarity to the forecast ensemble. This seems likely to improve out-of-sample performance but does not help with understanding the implicit assumptions driving the performance of multi-model, multi-team infectious disease forecast ensembles. As we have hypothesised that the use of absolute scoring measures is inappropriate and leads to performance characteristics that are unlikely to be favoured by forecast stakeholders more work should be done in this area. If new forecast ensemble methods are adopted as best practice by Forecast Hubs then follow-up work attempting to create surrogate forecast models should also use these approaches and this will likely alter the observed characteristics of the hub ensemble forecasts, for example, the tendency to be poorly calibrated. In September 2022, GitHub announced support for hosted GitHub Action runners with additional compute power (“GitHub Actions Larger Runners - Are Now in Public Beta” 2022). Whilst a paid feature this may allow more compute-intensive models, with fewer potential performance trade-offs, to be easily democratised though only if funds are available to support the hosting costs. One potential research area is to explore forecasting methods that can be used with a range of computing resources though this would require extensive evaluation and documentation to make it clear to users what the trade-offs between compute usage and forecast performance are. More work is needed to understand the best practice treatment of data revisions when evaluating forecasts and the potential bias these may cause. Lastly, here we have only explored a surrogate for an ensemble for a single disease, a limited set of locations, and a single target (incident cases), meaning our findings are difficult to generalise. Follow-up work should explore whether this behaviour holds across diseases, locations, and epidemiological targets where the behaviour of ensembles is notably different. However, this is limited to infectious diseases with similar large-scale forecast ensembling projects. These projects remain relatively rare despite them showing obvious promise to improve the forecasts available to stakeholders.

## CONCLUSIONS

We conclude that our simplified forecast model may have captured some of the dynamics of the hub ensemble but that more work needs to be done to understand the epidemiological model that represents its behaviour and whether or not this is the optimal choice for stakeholders’ requirements. We also conclude that our findings may be largely driven by the choice of evaluation measure used by the Forecast Hub. While this measure has desirable mathematical properties and is routinely used in a similar form e.g., in weather forecasting, it is subject to debate whether it appropriately reflects forecast users’ requirements and perceptions as to what makes a good forecast. Our work is useful for forecast users to understand the inherent assumptions of the forecasts they are making use of and to researchers thinking about how to develop forecasts that perform similarly to current multi-model and multi-team forecast ensembles that are trusted by stakeholders.

## Data Availability

All data and code are available here:

https://github.com/epiforecasts/simplified-forecaster-evaluation

https://doi.org/10.5281/zenodo.7189308

## ACKNOWLEDGMENTS

We thank the ECDC for supporting the forecasting hub, and all forecasters who submitted forecasts for making this study possible. We thank the Forecast Hub team for publishing all data in an accessible format. We thank the Epiforecast group for a productive discussion of an early version of this analysis. We thank Molly for being a good Labrador.

## ADDITIONAL INFORMATION AND DECLARATIONS

### Author contribution

SA conceived the study, developed the initial set of assumptions for the surrogate model, implemented the model into code, designed and conducted the forecast evaluation, and wrote the first draft of the manuscript. All other authors provided feedback on the manuscript and analyses and contributed to revisions. HG and KS reviewed the code and reproducibility of the analyses.

### Data availability

All data and code are available here: https://github.com/epiforecasts/simplified-forecaster-evaluation And are archived here:https://doi.org/10.5281/zenodo.7189308, https://doi.org/10.5281/zenodo.7189620

